# Early-life dentine-based elemental biodynamics and cord blood telomere length

**DOI:** 10.64898/2026.04.30.26351974

**Authors:** Bhargavi Srinath, Rohitha Ravisekar, Kshitij Sachdev, Joseph Eggers, Libni Avib Torres Olascoaga, Nia McRae, Inessa Lopez, Chelsea A. DeBolt, Aderonke Akinkugbe, Romana Ranchadiya, Martha M Téllez-Rojo, Chris Gennings, Robert B. Wallace, Robert Wright, Rosalind J. Wright, Manish Arora, Cecilia S. Alcala, Manasi Agrawal, Jamil M. Lane, Maria J. Rosa, Shoshannah I. Eggers, Vishal Midya

## Abstract

**Background:** Leukocyte telomere length (LTL) from cord blood is a marker of biological aging and long-term systemic health. Exposure to essential and toxic metals has been shown to influence LTL in a sexually dimorphic manner. However, little is known about the interplay between early-life longitudinal biodynamic patterns of these elements and cord blood LTL, as well as potential sex differences.

**Methods:** From an ongoing longitudinal birth cohort study in Mexico City, we used available tooth samples from 231 children (129 males and 102 females) to generate 16 elemental weekly time series of direct fetal intensities from the second trimester through four to five months after birth. We analyzed the dentine growth rings using Inductively Coupled Plasma Mass Spectrometry to generate time-resolved elemental intensities. The elements included were Li, Mg, Ca, Mn, Co, Ni, Cu, Zn, As, Sr, Mo, Cd, Sn, Ba, Pb, and Bi. LTL was measured in cord blood using qPCR. We used cross-recurrence quantification analysis and entropy-complexity-based measures to generate time-resolved features that quantify the synchronization of elemental biodynamics. A stability-selection approach using five-fold cross-validation of regularized ridge regression was used for feature selection, and covariate-adjusted linear models were used to estimate associations with LTL.

**Findings:** The biodynamic interaction of Mg-Co and Mn-Sn was identified as the most stable feature among male and female children, respectively. In males, higher vertical entropy (i.e., a measure of higher variability) of Mg-Co temporal biodynamics was associated with shorter LTL (β[95%CI]: -0.9[-0.14,-0.03]; p-value<0.01), but not in females (β[95%CI]:-0.02[-0.10,0.06]; p-value=0.60); whereas higher recurrence rate (i.e., a measure of higher synchronicity) of Mn-Sn temporal biodynamics was associated with longer LTL (β[95%CI]: 0.09[0.02,0.16]; p-value=0.01), in females but not in males (β[95%CI], 0.03[-0.04, 0.09]; p-value=0.39).

**Interpretation:** We demonstrate that time-varying multi-elemental synchronization of early-life elemental biodynamics, a potential marker of homeostatic balance, may be associated with cord blood-based telomere length in a sexual dimorphic manner.

## Introduction

Aging is a complex, heterogeneous process shaped by genetic predisposition, environmental exposures, and developmental determinants. [1, 2] Chronological age reflects time since birth, whereas biological age reflects the cumulative burden of cellular damage, environmental exposures, and shifts in homeostatic mechanisms.

[2] Several biomarkers have been proposed to predict biological aging, including DNA methylation age, telomere length, mitochondrial DNA content, and circulating analytes that show age-related change.[3] Among these, telomere length is one of the most widely studied as a potential marker of cellular aging and a long-term indicator of health trajectory. [4, 5] Telomeres are repetitive nucleoprotein structures at chromosome ends that shorten with each cellular replication cycle and in response to oxidative stress, eventually contributing to impaired tissue repair, reduced regenerative capacity, and cellular senescence. [6-9] Telomere dynamics begin in utero and are affected by maternal and early-life exposures. Cord blood-based leukocyte telomere length (LTL) at birth can therefore serve as an early indicator of biological aging. [10, 11] Shorter LTL has been associated with adverse developmental outcomes and health issues later in life in some populations, and newborn telomere length strongly predicts telomere length later in life, underscoring the importance of understanding its prenatal determinants. [12, 13]

Exposure to essential and toxic elements during fetal development represents one such determinant. Essential elements (e.g., zinc and copper) support antioxidant defense and DNA repair, whereas toxic metals (e.g., lead, cadmium, and arsenic) promote oxidative stress and genomic instability. [14, 15] Prenatal exposure to toxic metal mixtures was inversely associated with newborn LTL, and higher maternal antioxidant intake attenuated these effects.[16, 17] Furthermore, both metal toxicity and biological aging exhibit marked sexual dimorphism. [5] Telomere biology is sex-dependent: females have longer telomeres than males from birth onward, a difference that parallels the sex gap in life expectancy. [18]

However, not much is known about the interplay of elemental homeostasis (i.e., dynamic regulation of essential minerals and ions over time)[19] and LTL. Human regulatory mechanisms tightly control the dynamics of elemental homeostasis, and furthermore, exposure to external environmental perturbations (particularly in the form of environmental exposures) can significantly dysregulate homeostatic balance. [20] We hypothesize that the longitudinal dynamic patterns of elemental uptake levels (henceforth referred to as elemental biodynamics) may serve as a marker of homeostatic alterations.[21] While many biological processes (e.g., sleep patterns) have temporal biodynamic rhythms, fluid-based biomarkers of homeostasis reflect only a snapshot in time and capture limited information. [21, 22] Measuring biodynamic patterns of elemental concentrations from repeated blood samples is not feasible due to the need for frequent sampling, limited temporal resolution, and participant burden.

Calcified tissues such as teeth mineralize incrementally and then remain metabolically stable, incorporating elements during defined developmental windows and thus acting as archives of early-life exposure. [21] Laser ablation inductively coupled plasma mass spectrometry (LA-ICP-MS) applied to dentine growth increments allows retrospective reconstruction of elemental exposure histories from the second trimester through early childhood. [23] Tooth-based exposure assessment have been linked to neurological disregulation.[24] Given that telomere shortening is driven by oxidative and inflammatory stress, we hypothesize that early-life synchronization of elemental biodynamics may be associated with longer LTL. Although plausible explanations can be offered to link telomere shortening to early-life environmental exposures, there is little epidemiological evidence to support them. Therefore, in this study, we aim to investigate the interplay between early-life elemental biodynamics and cord blood telomere length (and any potential sexual dimorphic effect) using data from a longitudinal birth cohort. Figure 1 represents an overall schematic of the study.

**Figure 1:**
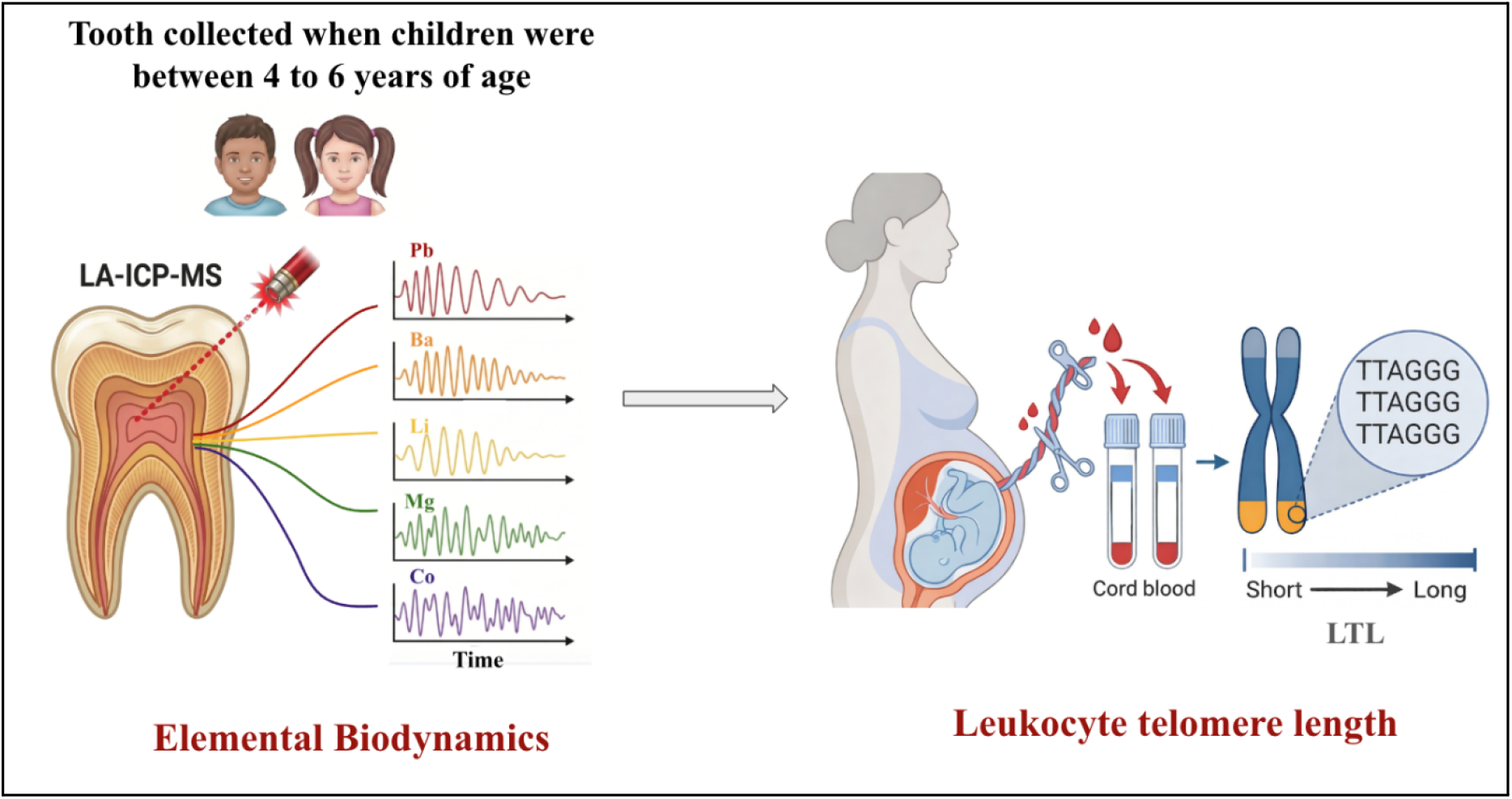
Overall Schematic of the study showing interplay of dentine-based early-life elemental biodynamics and cord blood leukocyte telomere length. LTL: Leukocyte Telomere Length

## Materials and Methodology

### Study Design and Population

This study prospectively included mother-newborn pairs from the Programming Research in Obesity, Growth, Environment and Social Stressors (PROGRESS) study, a longitudinal birth cohort that recruited participants between December 2007 and July 2011. [25] The study originally enrolled pregnant women who gave birth through the Mexican Social Security System. The eligibility criteria were: women from Mexico City with conceptuses at gestational age less than 20 weeks, age at pregnancy above 18 years, expectation to live in Mexico City for at least the subsequent 3 years, completed primary education, without a medical history of heart or kidney diseases, and did not consume alcohol daily nor use any steroid or anti-epilepsy medications. At enrollment and follow-up visits (second trimester, third trimester, and delivery), participant information was collected via questionnaire, including maternal age, smoking status, environmental smoking status, alcohol consumption, educational level, marital status, parity, and socioeconomic status. Women enrolled in this cohort voluntarily provided written informed consent. For the present analysis, we included 231 children (129 males, 102 females) who had both cord blood collected at delivery for LTL quantification and an extant deciduous tooth available for dentine-based elemental analysis. Deciduous teeth were obtained at ages 4-6 years upon natural exfoliation. This study was approved by the institutional review boards at the Icahn School of Medicine at Mount Sinai and the Mexican National Institute of Public Health. The participant flow from the original PROGRESS cohort to the final analytic sample is presented in Supplemental Figure S1.

### Dentine metal analysis

Metal concentrations in deciduous tooth dentine are well-validated biomarkers of early-life exposure and have been used to study neurologic and psychiatric outcomes.[25] In PROGRESS, shed deciduous teeth were collected by field staff during study visits or at home. The field staff registered the storage conditions reported by the mothers, noting whether the teeth were kept in a dry place. Envelopes were provided to participants to facilitate storage until a scheduled home pickup visit or directly when donated during the study visit. Teeth with caries, substantial loss of dentine or enamel, metal restorations, or developmental defects were excluded from analysis to avoid bias.[26] Teeth were washed and sectioned in a vertical plane. The neonatal line was identified to demarcate prenatal and postnatal developmental zones. Age along the crown was estimated from visible daily growth increments.[27] Elemental concentrations, including Li, Mg, Ca, Mn, Co, Ni, Cu, Zn, As, Mo, Sr, Cd, Sn, Ba, Pb, and Bi, were measured at 35 μm resolution by laser ablation inductively coupled plasma mass spectrometry (LA-ICP-MS). [28] The elemental intensities were recorded on a weekly basis starting from around the second trimester (16 weeks before birth) to about four to five months after birth. However, to maximize sample size given variable numbers of weeks captured per tooth, the statistical analyses were restricted to a common window from 16 weeks pre-birth to 16 weeks post-birth. Metals were normalized to calcium (^43^Ca) and corrected to the external standard NIST 610 to account for differences in mineral density between samples and instrumental drift. [29, 30] This yielded time-resolved elemental intensity signals for each element over the selected developmental window. Note that, for this analysis, we included both prenatal and postnatal dentine measurements. We justify this inclusion because there is no evidence of an abrupt, significant change in elemental biodynamics from the prenatal to the postnatal period, particularly in the first few months after birth. See the sensitivity analysis for additional comments and results.

### Cord blood collection and Telomere length analysis

DNA extraction, normalization, and qPCR-based measurement of LTL followed the methods described in detail previously. [31] Blood samples were taken from the venous umbilical cord blood during delivery. Samples were extracted using phenol-chloroform methods after red cell lysis and stored at 4°C. Excluding samples with insufficient volume or low DNA concentrations, samples were used to measure LTL using a modified qPCR method. [22, 32, 33] DNA was normalized to 2 ng/μL; concentration was confirmed with PicoGreen. Telomere primers were Telc 5′-TGT TAG GTA TCC CTA TCC CTA TCC CTA TCC CTA TCC CTA ACA-3′ and Telg 5′-ACA CTA AGG TTT GGG TTT GGG TTT GGG TTT GGG TTA GTG T-3′. The assay compares amplification of the telomere repeat relative to the single-copy albumin gene, yielding a telomere-to-single-copy gene ratio (T/S ratio). [21, 22] Reactions included antibody-mediated hot-start iTaq DNA polymerase, iQ SYBR Green Supermix, and a fluorescein passive reference dye. Thermal cycling began with 95 °C for 3 min to activate the hot-start enzyme, followed by 2 cycles of 15 s at 94 °C and 15 s at 49 °C, then 32 cycles of 15 s at 94 °C, 10 s at 62 °C, 10 s at 74 °C with signal acquisition, 10 s at 84 °C, and 10 s at 88 °C with signal acquisition. [21] A melting curve from 72 °C to 95 °C (0.5 °C per step) was run to verify amplification specificity. All samples were run in triplicate. A pooled quality-control sample (equal mass of DNA from all samples), normalized to 2 ng/μL, was included on each plate. Samples were distributed across six batches; each batch included a study-specific standard curve prepared from pooled DNA normalized to 30 ng/μL.[21] The T/S ratio (Telomere/single-copy gene ratio) was estimated using Bio-Rad software as the ratio of the starting quantities of telomere and albumin copy numbers, based on the standard curve prepared (Cq = slope × log10(Sq) + intercept). CV (Coefficient of Variation) was calculated for samples in triplicate. Samples with CV outside inter-quartile range were re-run, and the second LTL measure was recorded. If both had CV outside the threshold range, the sample was flagged as potentially unreliable. Samples with CV outside acceptable limits after multiple runs were excluded from further analysis.[21]

### Covariates

All regression models were adjusted for a common set of covariates based on prior literature and directed acyclic graphs.[23] The covariates included childhood factors like, gestational age at birth, Fenton Z-score (growth calculator for preterm infants)[34]; maternal and pregnancy factors like, maternal age during pregnancy, pre-pregnancy body mass index (BMI), education, socioeconomic status, smoking inside the home during pregnancy; hematologic factors like child hemoglobin and leukocyte counts to account for variability in telomere length; and factors related to quality control, batches of the assay and whether the sample was rerun (i.e. whether data comes from second LTL measurement).

### Statistics

#### Time-varying feature estimation

The statistical analysis was conducted in multiple stages. We extracted features that captured the time-varying biodynamics of elemental intensities. In particular, for individual elemental biodynamics, we calculated an univariate entropy-complexity measure [35]; whereas to explore pairwise bivariate elemental biodynamics, we used cross recurrence quantification analysis (CRQA). [25] Briefly, entropy-complexity measures estimate fluctuations in uncertain variability (i.e., irregularity) and the complexity of time-varying elemental intensities. For each elemental intensity, we estimated five features, namely the permutation entropy, spectral entropy, sample entropy, Hjorth mobility, and complexity. See further details in the Python Package AntroPy. [36] Multiple previous studies on elemental biodynamics have shown significant synergistic effects when pairwise bivariate elements are considered, relative to univariate elemental features. [37, 38] Therefore, our main analysis focused on bivariate elemental features. CRQA quantifies several aspects of dynamism through their co-occurrence behaviors. Moreover, it captures the dynamic coupling pattern over several lags to construct a two-dimensional map of trajectories, or, in other words, a map of phase space. [25, 39] Multiple features were then estimated from those two-dimensional maps. Given the fluctuations in recurrence sensitivities, 13 recurrence measures were computed over a range of radii to obtain robust estimates. [40, 41] For each time series, false nearest neighbors and mutual information were used to optimize the embedding dimension and time delay parameters, a standard approach in studies of nonlinear analysis. [26]

The measures captured from CRQA were: (1) Recurrence rate: represents the density of recurrence points in a Cross-Recurrence Plot, denoting the percentage of time two time-varying intensities are in a similar state. (2) Determinism: indicates strong coupling behavior or a similar structural pattern between the two time-varying intensities. (3) Average diagonal line length: estimates the average length of diagonal lines in the recurrence plot, signifying a more predictable system. (4) Longest diagonal line length: indicates a more stable or predictable shared dynamics between two time-varying intensities. (5) Divergence: it is defined as the inverse of the longest diagonal line in a recurrence plot. It signifies the rate of trajectory divergence, with greater divergence indicating weaker predictive dynamics. (6) Entropy diagonal lines: this indicates that the diagonal lines have a varied distribution of lengths, signifying high variability, lower regularity, or non-repeating interactions. (7) Laminarity: it is defined as the ratio of recurrence points forming vertical lines to the total number of recurrence points. High laminarity indicates stationarity or shared stability between two dynamic systems. (8) Trapping time: it calculates the average of the vertical lines in a cross-recurrence plot when two time-varying intensities are within a specific radius. (9) Longest vertical line length: A higher value indicates that the two time-varying intensities remained in a stable state for a longer period. (10) Entropy vertical lines: it measures the Shannon entropy of the histogram of vertical line lengths, which represents high complexity or irregularity during the stable periods of the time-varying intensities. (11) Average white vertical line length: it measures the average duration of time between two consecutive occurrences of similar states between two time-varying intensities. (12) Longest white vertical line length: it measures the maximum duration the two dynamic systems stay in a non-recurrent state, indicating the longest time gap between similarities in two time-varying intensities. (13) Entropy white vertical lines: it measures the Shannon entropy of the frequency distribution of white vertical line lengths, with a higher value indicating random, unpredictable patterns.

The Python library PyRQA was used for CRQA implementation, and the NoLiTSA library was used to optimize the embedding dimension and time delay parameters. [42] In total, we created 78 features from univariate entropy-complexity and 1687 from bivariate CRQA. All features were constructed in an outcome-agnostic way.

### Stability-based feature selection and Regression-based association

We conducted a feature selection stage because the number of bivariate biodynamic features greatly exceeded the sample size, thereby increasing the risk of false discoveries. Regularized ridge regression models were used and implemented through five-fold cross-validation. To ensure robustness and stability of the feature selection procedure, the ridge regression was repeated 100 times over randomly chosen seeds. In each iteration, the biodynamic feature with the largest absolute coefficient was noted. The biodynamic feature that most frequently occurred and had the highest absolute coefficient across the 100 iterations was selected for the next stage of regression-based association analysis. Among multiple types, this is a stability-selection method based on resampling techniques. [43] Note that such a choice of a single “top” feature with the highest absolute coefficient did not necessarily imply statistical significance in the regression-based analysis. This is because a feature was selected at each iteration, regardless of AUC or other performance metrics. This choice of top feature ensures the minimization of false discoveries, since at each iteration, there will always be a feature with the maximum absolute coefficient. Next, this chosen feature is used in a regression-based inference model that adjusts for covariates and confounders. The regression models were also adjusted for the mean intensity of the elements whose elemental biodynamic feature was considered for modeling. To minimize the influence of outliers, the biodynamic features were converted to deciles. Analyses were conducted separately for each sex to account for potential sexual dimorphism.

Finally, we repeated the entire feature-selection and regression-based pipeline using the univariate entropy-complexity features. There were very few missing values in the covariates (<10%), and no missingness was observed in the biodynamic features. Feature selection and regression-based modeling were conducted in R (version 4.5.1).

### Sensitivity analyses

To evaluate the robustness of our findings, we performed several sensitivity analyses. (1) The main bivariate pair-wise analysis was repeated by converting the outcome into deciles to ensure the directionality of the results is not affected by outliers in the outcome. (2) For the univariate features as well, we converted the LTL into deciles and repeated the analysis to ensure the directionality does not alter due to outliers. (3) We estimated distribution-free robust randomization-based p-values for each of the main associations to ensure that model-based p-values are not significantly different. [28] (4) And lastly, to ensure that elemental biodynamics do not drastically alter between the prenatal period and just after a few months after birth, we considered repeating the main analyses, constraining the elemental biodynamics to only the prenatal period.

## Results

The demographic details are presented in Table 1. Among the 231 samples, there were more male children than female (55.8% and 44.2%, respectively). The mean maternal age at pregnancy was 28.16 ± 5.55 years. Mean pre-pregnancy BMI was 26.30 ± 4.19kg/m^2^. The majority of the women were from lower socioeconomic levels (55.4%). Only about a quarter (24.68%) of mothers had a high school education or higher. About a third (30.57%) of the mothers reported second-hand smoke exposure. The mean cord blood hemoglobin from the extracted blood was 15.79 ± 1.85 g/dL, with no significant difference between male and female children. Mean count of cord blood leukocytes was higher in female children (12.08 ± 4.05 10^3^/ml). The relative LTL in the cord blood was slightly higher in the males (1.27 ± 1.89). There were no significant differences in mean elemental intensities by tooth type. The elemental mean intensities between males and females were not significantly different (Supplementary Table 1).

**Table 1:** Overall participant characteristics.

| **Variables** | **Overall**<br>**N = 231** | **Male**<br>**N = 129**<br>**(55.8%)** | **Female**<br>**N = 102**<br>**(44.2%)** | **p-value<sup>c</sup>** |
| --- | --- | --- | --- | --- |
| ***Maternal age (in years), Mean (SD)*** | 28.16 (5.55) | 28.43 (5.26) | 27.83 (5.91) | 0.31 |
| ***Maternal pre-pregnancy BMI, Mean (SD)<sup>a</sup>*** | 26.30 (4.19) | 26.87 (3.99) | 25.57 (4.35) | 0.004 |
| ***Maternal Education level, n(%)*** |  |  |  |  |
| Primary& Secondary level | 83 (35.93%) | 56 (43.41%) | 27 (26.47%) | 0.02 |
| Post-secondary level | 91 (39.39%) | 43 (33.33%) | 48 (47.06%) |  |
| University level | 57 (24.68%) | 30 (23.26%) | 27 (26.47%) |  |
| ***Maternal SES, n(%)<sup>b</sup>*** |  |  |  |  |
| Lower | 128 (55.41%) | 73 (56.59%) | 55 (53.92%) | 0.92 |
| Middle | 85 (36.80%) | 46 (35.66%) | 39 (38.24%) |  |
| Upper | 18 (7.79%) | 10 (7.75%) | 8 (7.84%) |  |
| ***Maternal Second-hand smoke exposure, n(%)*** | 70 (30.57%) | 35 (27.56%) | 35 (34.31%) | 0.27 |
| ***Cord blood hemoglobin(g/dl), Mean (SD)*** | 15.79 (1.85) | 15.92 (2.06) | 15.61 (1.55) | 0.24 |
| ***Cord blood leukocytes($10^3$/ml), Mean (SD)*** | 12.09 (3.77) | 11.51 (3.44) | 12.83 (4.05) | 0.01 |
| ***LTL in cord blood, Mean (SD)*** | 1.27 (1.89) | 1.35 (2.50) | 1.16 (0.50) | 0.54 |
| ***Fenton Z-score, Mean (SD)*** | -0.41 (0.80) | -0.43 (0.86) | -0.39 (0.73) | 0.61 |

<sup>a</sup>SES, socioeconomic status; <sup>b</sup>BMI, body mass index; SD, standard deviation;
<sup>c</sup>p-values were obtained using Pearson's Chi-Squared Test or Wilcoxon rank sum test as required.

### Illustration of elemental biodynamic features

Prior to presenting the main results, we present a visual illustrative example of time-varying biodynamics for two elements, Co and Mg, in a male child with a longer telomere length and another male child with a relatively shorter one. The purpose of this illustration is to demonstrate how synchronization (and asynchronization) of two time-varying intensities implies. Longer (and shorter) telomere length is defined based on the median for ease of interpretation. We presented the time-varying illustration in Figure 2. The temporality of Co and Mg dentine-intensities showed a synchronous (albeit time-lagged) and a relatively asynchronous pattern in two male children with longer or shorter telomere lengths, respectively (Figure 2A and 2B). Moreover, in Figures 2C and 2D, we presented the corresponding cross-recurrence plots, which showed regions of closeness between the temporal intensities, as well as regions where the two intensities were far apart. The recurrence plots visualized the dynamic trajectory of elemental intensities, quantifying the time to return to a previous value (or to a neighborhood within a radius) of the intensities. Multiple temporal features were calculated from these recurrence plots, revealing significant differences. In total, 78 and 1687 univariate entropy-complexity and pair-wise CRQA features were calculated.

**Figure 2:**
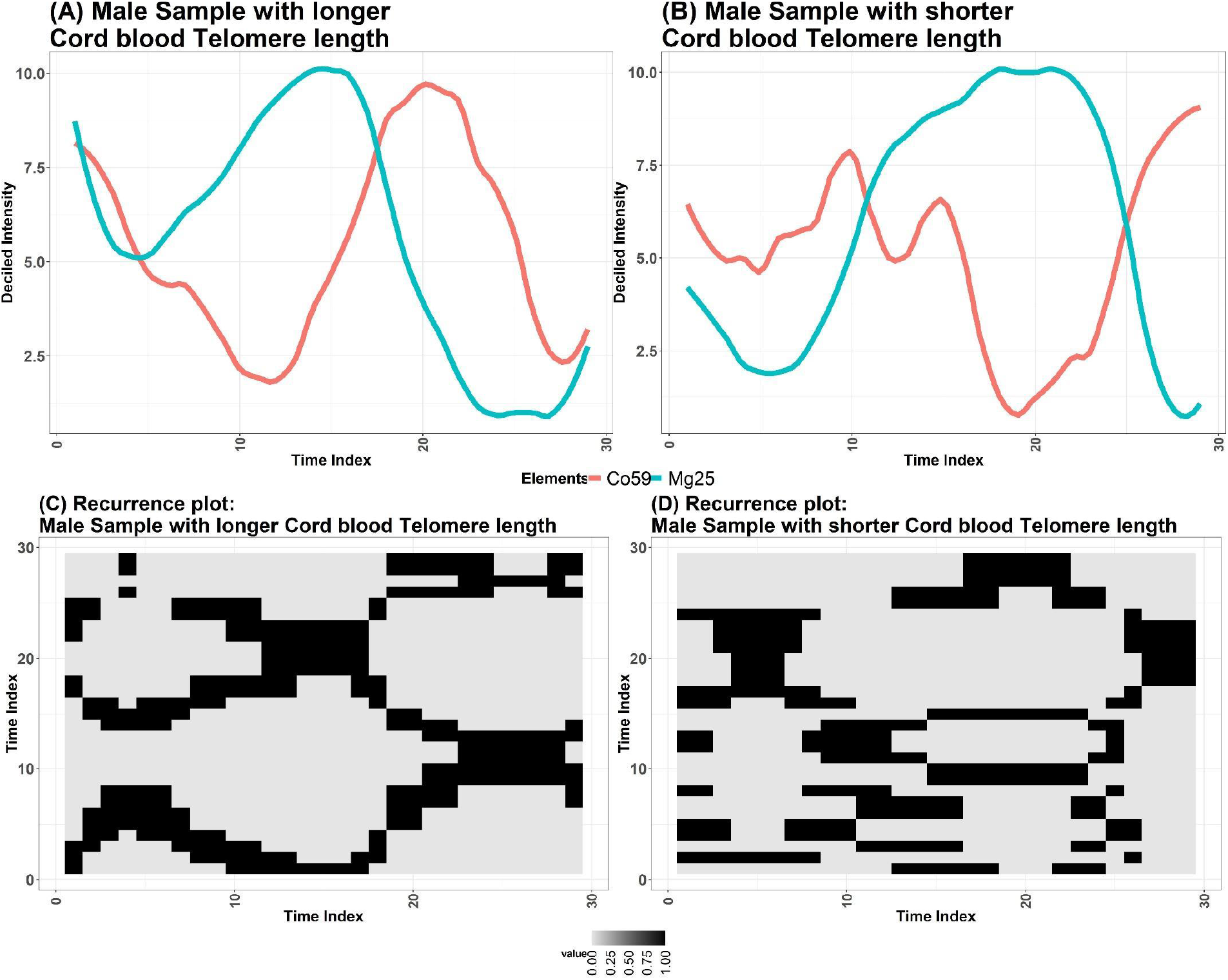
A visual illustration of elemental biodynamics of Mg-Co intensity (and cross-recurrence structure) in a male child with longer versus another male child with shorter cord blood telomere length. Longer (and shorter) telomere length is defined based on the median for ease of interpretation. Panels A and B show deciled Mg and Co dentine intensity trajectories over the perinatal window for a male child with longer LTL and a male child with shorter LTL. Panels C and D display the corresponding cross-recurrence plots for Mg-Co in these same children, illustrating denser, more structured recurrence in the child with the longer LTL (2C) compared with the child with the shorter LTL (2D).

### Elemental biodynamics and elemental intensities are not correlated

Figure 3 highlights the correlation between biodynamic features and mean intensities. The main CRQA feature, vertical entropy of Mg-Co, was weakly correlated with both mean Mg intensity (ρ = 0.11) and mean Co intensity (ρ = 0.13) (Figure 3A). In Figure 3B, we show an analogous correlation matrix for Mn-Sn recurrence rate and mean Mn and Sn intensities in females. Mn-Sn recurrence was weakly and inversely correlated with mean Mn intensity (ρ = -0.16) and mean Sn intensity (ρ = -0.06). These correlations indicate that the biodynamic feature possibly captures temporal information largely independent of overall elemental intensity. These mean elemental intensities were included as covariates in the regression models to ensure that the CRQA-derived biodynamic features provide information about temporal structure and pairwise synchronization that is not redundant with mean elemental intensity levels.

**Figure 3:**
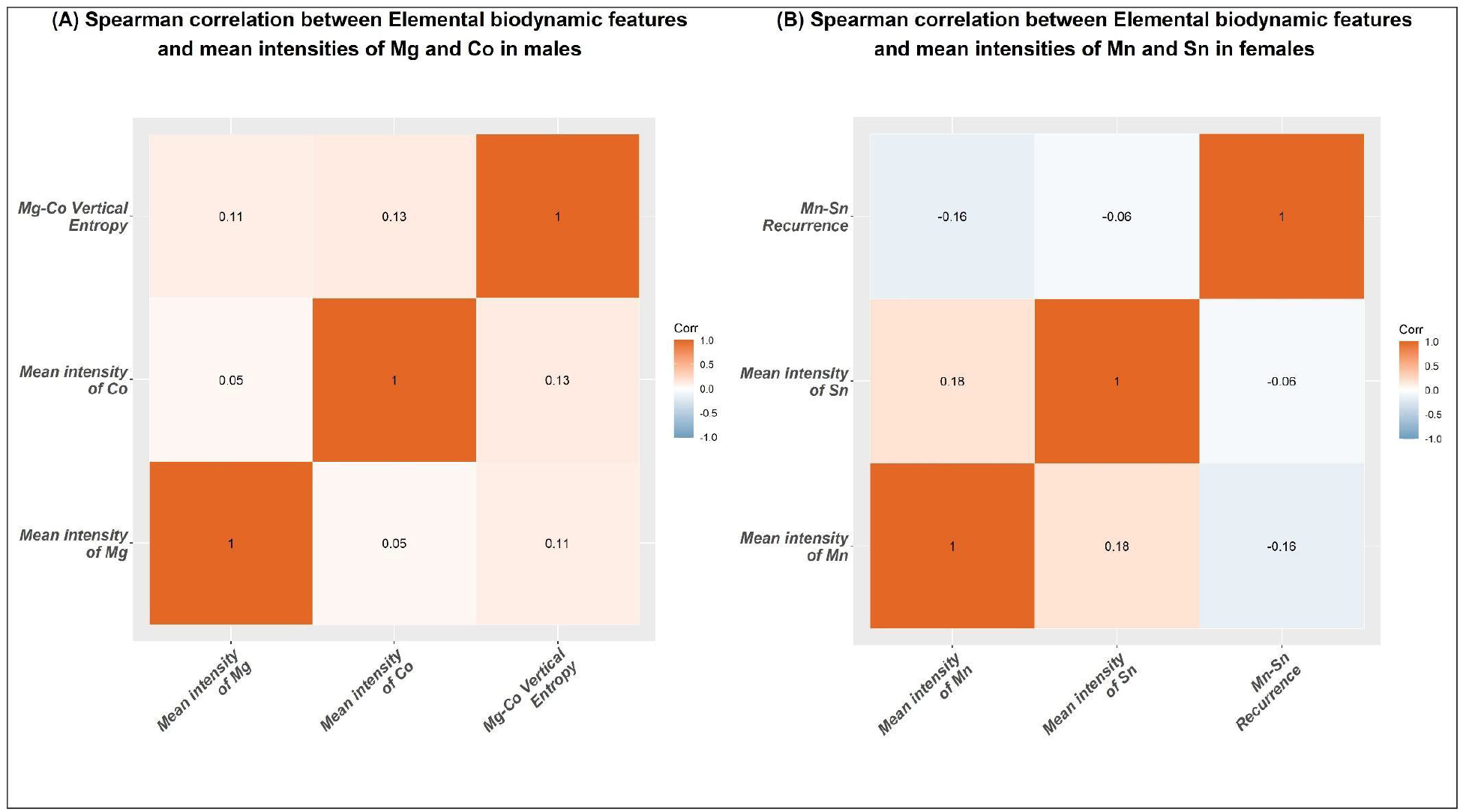
Spearman correlation matrices between the selected cross-recurrence quantification analysis features and their corresponding mean elemental intensities, stratified by sex. (A) shows correlations among Mg-Co vertical entropy and the mean dentine intensities of Mg and Co in males. (B) shows correlations among a Mn-Sn recurrence measure and the mean dentine intensities of Mn and Sn in females.

### Sex-specific associations for biodynamic features

In male children, across 100 five-fold cross-validated replications, only two top biodynamic features (with the largest absolute coefficients) were consistently chosen: vertical entropy of Mg-Co and laminarity of Mg-Co, occurring 65% and 35% of the time, respectively. In females, Mn-Sn recurrence was the only top feature that occurred across all the repetitions. This indicates that the most frequent time-resolved features differed between the two sexes. When the procedure was applied to the univariate entropy-complexity feature set, permutation entropy for Barium and spectral entropy of Zinc were chosen as the top features. Since only the top feature was chosen in each analysis, the issue of false discoveries was minimized. Next, these top features were used in a regression with a priori chosen covariates. The biodynamic features were converted into deciles. The results of the regression analysis were presented in Figure 4. Higher Mg-Co vertical entropy was associated with lower LTL in males (β[95% CI]: -0.09[-0.14, -0.03]; p=0.004) but not in females (β[95% CI]: -0.02[-0.10, 0.06]; p=0.60). The regression with Mg-Co vertical entropy was adjusted for the mean elemental intensities of Mg and Co. Higher Mn–Sn recurrence was associated with higher LTL in females (β[95% CI]: 0.09[0.02, 0.16]; p=0.01) but not in males (β[95% CI]:0.03[-0.04, 0.09];p=0.39). Similar to the regression in males, the regression with Mn–Sn recurrence was adjusted for the mean elemental intensities of Mn and Sn. These estimates are presented as a forest plot in Figure 4A. To further illustrate the robustness of the associations and to visually represent the differences in LTL distribution across the chosen biodynamic features, boxplots of LTL were presented for dichotomized biodynamic features in Figures 4B and 4C. The results of the univariate entropy-complexity features were presented in Supplementary Table 2. In males, higher permutation entropy of Ba was associated with shorter LTL (β[95% CI]:-0.05[-0.11, 0.02];p=0.15), and in females, spectral entropy of Zn was associated with longer LTL (β[95% CI]:0.06[-0.01, 0.13];p=0.12). Note that we presented results based only on the chosen stable features to ensure strict minimization of false discoveries.

**Figure 4:**
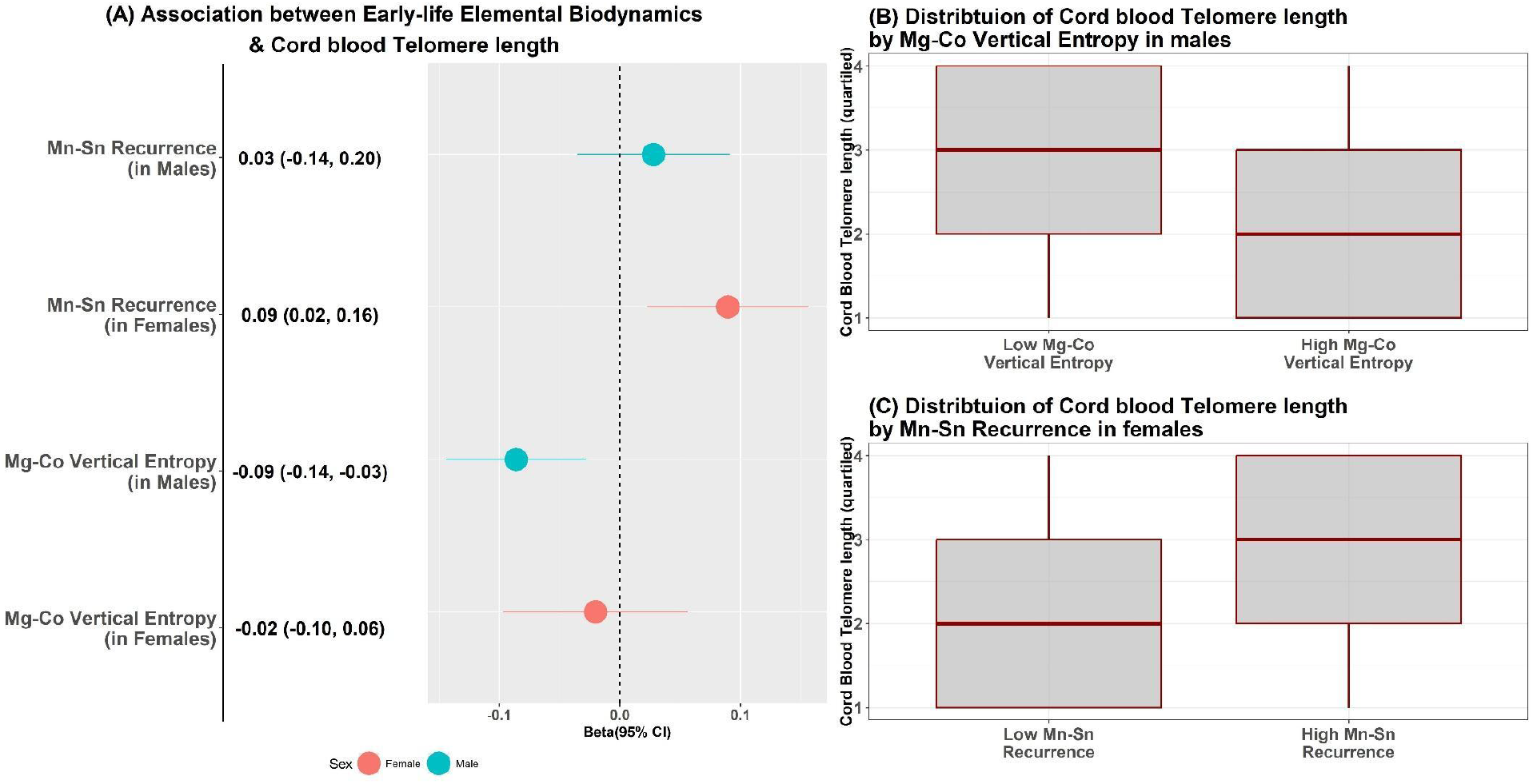
Sex-stratified associations between pair-wise elemental biodynamic features and cord blood LTL. (A) Forest plot showing covariate-adjusted β-coefficients and 95% confidence intervals from regression models for two CRQA features, Mg-Co vertical entropy and Mn-Sn recurrence rate in males and females, respectively. Figure 4B and 4C illustrate the corresponding boxplots (distributions) of LTL across low versus high (with respect to median) Mg-Co vertical entropy (males) and Mn-Sn recurrence (females).

Sensitivity analyses showed that (1) even after converting the LTL outcome into deciles, the directionality of the main CRQA-based pair-wise associations remained unaltered, showcasing the robustness of our result (see Supplementary Figure S1). Furthermore, although the main univariate associations were not statistically significant, regression with the LTL outcome converted to deciles showed that the directionality remained similar to the main associations (see Supplementary Table 2). In summary, we illustrated that our modeling techniques and results were minimally affected by outliers in LTL. (3) We estimated distribution-free robust randomization-based p-values for each of the main associations. To estimate the permutation p-values, we iterated the process 10^5^ times. In males, for the Mg-Co vertical entropy and Mn-Sn recurrence rate, the robust p-values were below 10^-5,^ and 0.43, respectively. In females, the robust p-values for the Mn-Sn recurrence rate and Mg-Co vertical entropy were also below 10^-5^ and 0.57, respectively (see Supplementary Table 3). (4) Lastly, we repeated the main analyses with Mg-Co and Mn-Sn elemental biodynamics restricting only to the prenatal period (shorter time series with respect to the whole early-life prenatal and postnatal periods). As hypothesized, the directionality of the associations remains unaltered between the prenatal and early-life periods, possibly indicating that elemental biodynamic patterns do not change substantially immediately after birth. For males, the beta estimate for only prenatal Mg-Co vertical entropy did not differ significantly from that for the overall early-life period (<10%). For females, the beta estimate for only prenatal Mn-Sn recurrence rate differed by 40% from that for the overall early-life period. This might have been due to a relatively short time series; to accurately estimate stable biodynamics—defined as the underlying long-term structural patterns or equilibrium behaviors—a significantly longer time series is generally required than for short-term forecasting. [44, 45]

## Discussion

We integrated high-resolution, temporally dense weekly direct fetal elemental intensity data to demonstrate that systemic dysregulation of elemental biodynamics is associated with cord blood LTL and to illustrate that time-varying elemental biodynamics may have possible sexual dimorphic associations with LTL. Mg-Co vertical entropy emerged as the dominant biodynamic feature in males, whereas Mn-Sn recurrence emerged in females, suggesting that homeostatic coupling among element pairs may relate to early-life telomere setting in a sex-dependent manner. These findings extend a growing literature linking elemental homeostasis and nutritional elements to oxidative stress, inflammation, and telomere biology, but emphasize dynamic co-regulation rather than a few time-averaged concentrations. Our results illustrate that early-life elemental biodynamics may serve as a predictive framework for an underlying physiological process that may predict systemic health.

Methodologically, tooth-matrix LA-ICP-MS provides a retrospective record of pre- and postnatal windows of direct fetal exposure, while the novel elemental biodynamic measure quantifies the synchronization and recurrence in coupled elemental trajectories. The construction of measures for elemental homeostasis requires high-frequency longitudinal data, but usual fluid-based biomarker data (urine, whole blood, or plasma) are often limited to a single or a few time points, mostly due to challenges in collection and storage. Dentine-based intensity data, therefore, provide a unique opportunity to study elemental homeostasis. The use of high-frequency intensity data, along with novel information-theory-based techniques to study the nonlinear dynamics of intensity, is unique to this study. Moreover, because newborn telomere length does not shift its relative position (within the TL distribution) in early childhood and young adulthood [31], this research provides initial evidence that early-life elemental homeostasis may even predict TL in early childhood.

Aging could be considered a progressive loss of homeostatic resilience. Disruptions in elemental biodynamics have been increasingly linked to phenotypes related to aging, particularly biomarkers such as telomere length.

[46] TL shortens progressively with each cell division, the process accelerated by oxidative stress and inflammation. [47] Macro-minerals such as Ca and Mg, and trace metals such as Mn, Cd, and Pb, participate in biochemical networks that sustain and regulate cellular function. Exposomic studies have demonstrated that metal patterns are not static. [48, 49] Elements act as enzyme cofactors, and their dysregulation can disrupt metabolic equilibrium and induce oxidative stress. [50, 51] Certain elements (e.g., Zn and Se) support antioxidant defense mechanisms, including the superoxide-dismutase and glutathione-peroxidase pathways. Others (Fe, Cu, and Mn) serve as redox-active metals and catalyze the formation of reactive oxygen species, which, in turn, damage DNA, proteins, and lipids, leading to systemic imbalance. [52-54]

Mg and Co participate in bioenergetic and cofactor networks [55]; Mg and Sn may reflect overlapping environmental sources or correlated metabolic handling in this context. [29, 30] Observed dimorphism is plausibly consistent with baseline sex differences in LTL, immune cell composition, and hormonal milieu, which can modify both elemental regulation and telomere maintenance; however, mechanistic inference remains limited in observational data. [56, 57] Mg and Co may interact antagonistically, with Mg acting as a protective agent against Co-induced cardiotoxicity and cellular damage. [58-60] However, little is known about the interaction between Mn and Sn. Mn homeostasis is heavily reliant on iron (Fe) levels, with high Mn inhibiting Fe absorption. [61-63] Studies have shown that Sn interferes with Fe uptake in intestinal mucosal cells. High intake of inorganic Sn has been shown to reduce Fe absorption. [64-66] Therefore, it is plausible that the elemental biodynamic interaction between Mn and Sn occurs by modulating Fe dynamics. Elemental homeostasis is essential for cellular redox balance, DNA repair capacity, mitochondrial function, and inflammatory and epigenetic regulation, mechanisms that overlap and greatly influence telomere length. Whether through nutritional deficiency, environmental exposures, or metabolic disease, disruption to the cellular homeostatic mechanisms of these elements contributes directly or indirectly to telomere shortening.

Sexual dimorphism in telomere length is well established. Females have been found to exhibit longer telomeres than males across the lifespan.[67] While hormones and genetic factors play major roles, the primary mechanism is differences in oxidative stress and immune function between the sexes. [68-70] In men, baseline oxidative and metabolic stress are innately higher, potentially accelerating telomere shortening.[71] Our results denote a drastic difference in the sex-specific effect sizes; however, the sex-specific confidence intervals do overlap-this might have been possibly due to relatively smaller sample sizes. Our results, therefore, may imply that sex-specific variation in element metabolism and distribution affects biodynamic patterns, which are likely to amplify or mitigate telomere attrition.

This study also has a few limitations. First, the study had a moderate sample size and lacked external validation. Given that we provided a proof-of-concept to explore the interplay between elemental biodynamics and cord-blood LTL, we plan to use these results to validate them in external cohorts for future studies. Second, we did not explore the underlying mechanisms of inflammation or oxidative stress, which might serve as mediators or moderators in the pathway linking elemental biodynamics and LTL. This will be extensively studied in our future work. Third, measurement errors in telomere length measurement and in dentine metal intensity estimation may have biased our results. However, we ensured that we adjusted for batch effects and any potential systematic deviations from the norm (outside the coefficient of variation) in our modeling approaches. For random errors, we would expect results toward the null, but our results consistently reproduced the main finding in sensitivity analyses. Fourth, this work presented results that are associative in nature and was conservative in its treatment of elemental biodynamic features. Given a moderate sample size, we adopted a rather cautious approach to ensure strict minimization of false discoveries. In future work with a larger sample size, we hope to use multiple biodynamic features to create predictive models of telomere length, which can then be used as an instrument for predicting biological aging. Our results should not be considered from a causal perspective, but rather from a predictive viewpoint, where one’s underlying elemental biodynamics may be able to inform systemic health. Strengths of this study include the novel use of objective tooth-based archives to obtain direct fetal elemental intensities, the application of information-theoretic feature extraction techniques of elemental biodynamics, a detailed exploration of sexual diamorphic effects of elemental homeostasis, and the potential exploration of a novel technique to quantify biological aging through the study of elemental biodynamics.

In conclusion, using time-resolved dentine-based direct fetal intensity and cord blood telomere length, we demonstrate that time-varying synchronization of early-life elemental biodynamics may be associated with cord blood-based telomere length in a sex-dependent manner. These results support elemental biodynamics as a complementary lens on early biological aging.

## Data sharing statement

Datasets generated and analyzed in the current study are not publicly available because they contain private patient health information. However, both codes and de-identified data could be made available on reasonable justification and subject to necessary clearances from the PROGRESS data team upon written request (with a proposal of how the data will be used) to Dr. Robert Wight.

## Acknowledgements

This study is supported by funding from the US National Institutes of Health (R21ES037112, P30ES023515, R01ES026033, U2CES030859, U2CES026561, R35ES030435, R01TS000324, and UL1TR004419)

