## Supplementary Material for "Early-life dentine-based elemental biodynamics and cord blood telomere length"

### Supplementary Figures

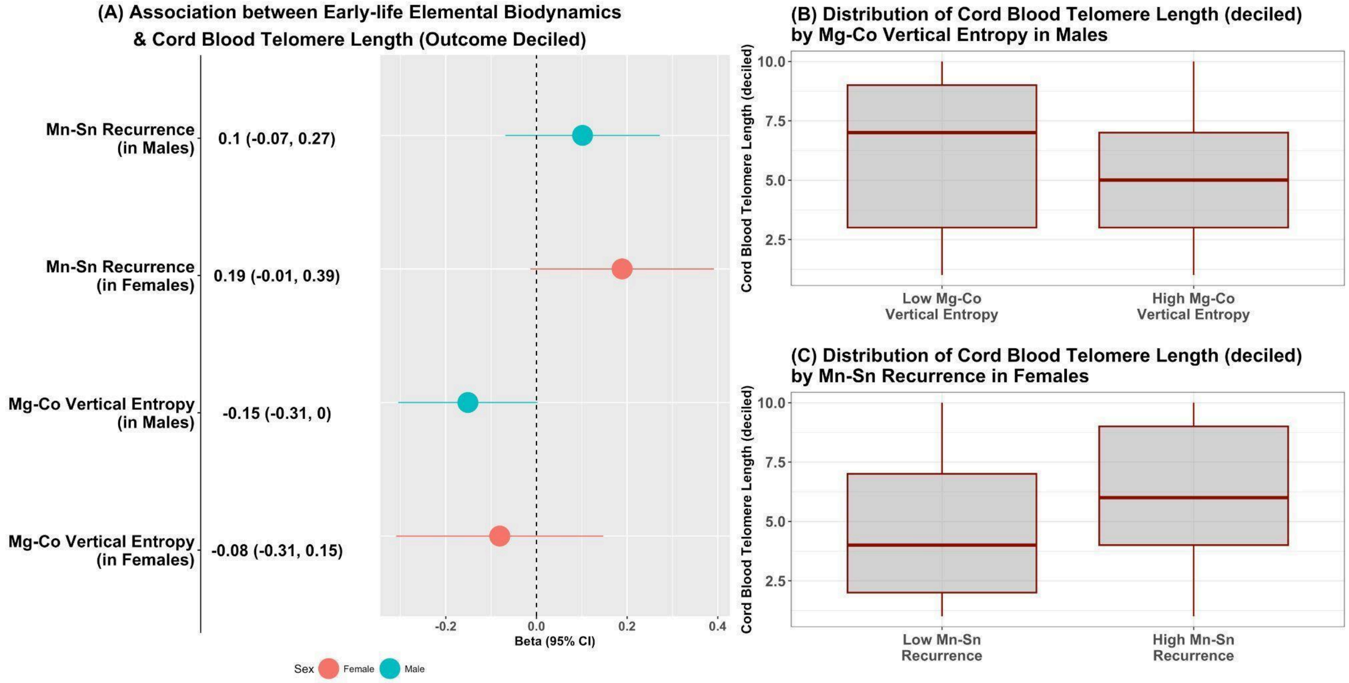


**Supplementary Figure S1: Deciled Cord blood telomere length (T/S ratio) and elemental biodynamic features, analysis stratified by sex.** (A) Forest plot (beta[95% CI]) of covariate-adjusted associations between early-life dentine-based elemental biodynamics and cord blood leukocyte telomere length (LTL) when the outcome was transformed into deciles. Associations were presented for bivariate cross-recurrence quantification analysis-based features, including Mn-Sn recurrence and Mg-Co vertical entropy. Analysis was stratified by sex. The vertical dashed line represents the null. (B) Among males, the distribution of deciled LTL comparing participants with low versus high Mg-Co vertical entropy (dichotomized at the median). (C) Among females, distribution of deciled LTL comparing participants with low versus high Mn–Sn recurrence (dichotomized at the median). Box plots show median (horizontal line), interquartile range (box), and whiskers to the most extreme points within 1.5*IQR.

### Supplementary Tables

**Supplementary table 1: Sex-stratified summary of tooth-dentine mean LA-ICP-MS elemental intensities**

| Mean Intensity (SD) | Male (N = 129) | Female (N = 102) | p-value |
|---|---|---|---|
| Li | 0.04 (0.93) | -0.05 (1.08) | 0.25 |
| Mg | 0.02 (1.06) | -0.02 (0.93) | 0.6 |
| Ca | 0.06 (1.04) | -0.08 (0.95) | 0.38 |
| Sr | 0.00 (0.98) | -0.01 (1.03) | 0.93 |
| Mn | -0.02 (0.97) | 0.03 (1.04) | 0.43 |
| Co | -0.01 (0.98) | 0.02 (1.03) | 0.48 |
| Ni | -0.02 (0.95) | 0.03 (1.06) | 0.51 |
| Cu | 0.01 (0.81) | -0.01 (1.2) | 0.67 |
| Zn | -0.07 (0.96) | 0.09 (1.05) | 0.26 |
| As | -0.12 (1.05) | 0.15 (0.92) | 0.07 |
| Mo | -0.02 (0.87) | 0.02 (1.15) | 0.66 |
| Cd | -0.07 (1.00) | 0.09 (0.99) | 0.13 |
| Sn | -0.04 (0.91) | 0.04 (1.1) | 0.87 |
| Ba | -0.03 (0.98) | 0.04 (1.02) | 0.59 |
| Pb | -0.05 (0.99) | 0.06 (1.02) | 0.25 |
| Bi | 0.01 (0.83) | -0.01 (1.18) | 0.98 |

Values are presented as Mean (SD). Mean intensities were calculated as the average laser ablation inductively coupled plasma mass spectrometry (LA-ICP-MS) signal per child across all weekly dentine measurements from approximately 16 weeks before birth to almost 16 weeks after birth. P-values were obtained from Wilcoxon rank-sum tests comparing males and females.

**Supplementary Table 2: Associations between univariate elemental biodynamic features (entropy-complexity) and cord blood leukocyte telomere length (LTL) at birth, stratified by sex.**

| | | **Males (n=129): permutation entropy of Ba138** | | **Females (n=102): Spectral entropy of Zn66** | |
|---|---|---|---|---|---|
| **Type of analysis** | **Model specification** | **β (95% CI)** | **p-value** | **β (95% CI)** | **p-value** |
| Main model | Continuous outcome-Deciled feature | -0.05 (-0.11, 0.02) | 0.15 | 0.06 (-0.01, 0.13) | 0.12 |
| Sensitivity analysis | Continuous outcome-Continuous feature | -0.18 (-0.36, -0.01)* | 0.04* | 0.17 (-0.04, 0.38) | 0.11 |
| Sensitivity analysis | Deciled outcome: Deciled feature | -0.10 (-0.26, 0.06) | 0.23 | 0.16 (-0.05, 0.38) | 0.14 |

*p<0.05. Models were adjusted for Fenton Z-score for BMI, maternal age, maternal pre-pregnancy BMI, smoking inside the home during pregnancy, hemoglobin, leukocyte count, maternal education, socioeconomic status, telomere assay plate, and telomere quality flag. All models were further adjusted for the log-transformed mean intensity of the corresponding element.

**Supplementary Table 3: Model-based p-value and robust permutation p-value for bivariate pairwise regression models.**

| CRQA feature | Model-based p-value | | Robust permutation test-based p-value | |
|---|---|---|---|---|
| | Male | Female | Male | Female |
| Mg-Co vertical entropy | 0.004 | 0.60 | $<10^{-5}$ | 0.57 |
| Mn–Sn recurrence | 0.39 | 0.01 | 0.43 | $<10^{-5}$ |

Distribution-free robust randomization-based p-values were estimated for each of the main associations. To estimate the permutation p-values, we iterated each regression $10^5$ times to obtain a stable estimate.
